# Race and Ethnic Disparities in Catheter Ablation Utilization for Atrial Fibrillation: A Meta-analysis and Framework for Implementation Research

**DOI:** 10.1101/2024.05.22.24307768

**Authors:** Waseem Nosair, Jamal Smith, Sarahfaye Dolman, Paul Kolm, Sung Lee, Apostolos Tsimploulis, Athanasios Thomaides, David Strouse, William S. Weintraub

## Abstract

**Background:** Race/ethnic disparities in catheter ablation utilization for atrial fibrillation (AF) have been reported in the literature, however the data have not been systematically reviewed.

**Objectives:** To perform a systematic review and meta-analysis of studies reporting on disparities in the utilization of catheter ablation (CA) and to explore possible root causes of disparities using a behavioral model of health service utilization.

**Methods:** We searched PubMed/MEDLINE, Web of Science and Embase for studies reporting on race/ethnic disparities in the utilization of CA for AF in the United States. A meta-analysis was performed on a subset of included articles using a random-effects model. Publication bias was assessed for race/ethnic groups pooled from 10 or more studies. We adapted a behavioral model of health service utilization to identify root causes of disparities.

**Results:** Our search identified 20 studies published between 2011 and 2023, representing 47,700,642 patients with AF of whom over 561,490 underwent CA. Compared to non-Hispanic White patients, racial minorities had a lower odds of utilization of catheter ablation: 0.68 (95%CI 0.58 – 0.77) for non-Hispanic black (NHB) patients, 0.72 (95%CI 0.65 – 0.79) for Hispanic/LatinX (HLx) patients, and 0.62 (95%CI 0.45 – 0.78) for Asian patients. Other race groups were excluded due to insufficient data. There was a moderate to high degree of between-study heterogeneity for each race/ethnicity group: HLx (I^2^ = 58.2%), Asian (I^2^ = 80.9%), and NHB (I^2^ = 90.4%). Only NHB patients had sufficient data to generate a funnel plot which showed evidence of publication bias.

**Conclusions:** The high between-study heterogeneity reveals varying degrees of disparities across studies and settings. Further research adjusting for patient-provider preferences and factors, echocardiographic data and social determinants of health is needed to clarify root causes of disparities and to promote equitable adoption of this important therapy in AF care.

## Introduction

Atrial fibrillation (AF), the most common arrhythmia, is associated with increased morbidity, mortality, and health care costs (1). Racial and ethnic disparities in AF diagnosis and management have been documented but remain poorly understood. While non-Hispanic White (NHW) patients have a higher incidence and prevalence of AF, racial and ethnic minorities have a higher morbidity and mortality (2,3). In the Outcomes Registry for Better Informed Treatment of Atrial Fibrillation (ORBIT-AF) study, for example, non-Hispanic black (NHB) patients were more likely to report severe symptoms and lower quality-of-life scores(4). In the Atherosclerosis Risk in Communities (ARIC) study, NHB compared to NHW patients, had a 1.5 to 2 times higher risk of stroke, heart failure, coronary heart disease, and mortality (5). Similar findings have been noted among other race and ethnic groups (3).

The management of AF has evolved over time, with a recent shift towards early rhythm control to prevent disease progression and reduce adverse cardiovascular outcomes (6). Race and ethnic disparities in the utilization of rhythm control, including the use of electrical cardioversion, anti-arrhythmic drugs (AAD) and surgical or catheter ablation (CA), have been reported in several observational studies. However, the data have not been systematically reviewed and analyzed. The present paper seeks to systematically review inequities in the utilization of rhythm control, specifically CA. This is particularly timely and relevant, given the recent promotion of CA to a first-line indication for atrial fibrillation in appropriately selected patients in national guidelines (7). We adapted a behavioral model of health services utilization to identify factors that may perpetuate race/ethnic disparities in the utilization of this important therapy in the United States and to highlight areas for future research and interventions.

While individual articles identified in our reviews may define race or ethnicity differently, we use the terms interchangeably, recognizing that they are purely social constructs rather than biologic. Race refers to the social construction and categorization of people based on perceived shared physical traits and to a lesser degree culture, whereas ethnicity characterizes people based on having shared culture, history, and common ancestry. The term LatinX is included as a gender-neutral term used to refer to people of Latin American cultural or ethnic identity in the United States. The gender-neutral (-x) suffix replaces the (-o/-a) ending of Latino and Latina that are typical of grammatical gender in Spanish. We acknowledge that such social constructs have perpetuated bias and result in the maintenance of sociopolitical hierarchy; however, we find that as self-reported data they also allow the identification of marginalized groups that may underutilize catheter ablation.

## Methods

### Search Strategy

To investigate the association between race/ethnicity and utilization of CA for AF rhythm control, we performed a systematic review and meta-analysis, following PRISMA guidelines, of observational studies identified through the literature search. The PRISMA checklist is attached (Supplement 1). The literature search focused on publications reporting associations between race/ethnicity and the utilization of CA and rhythm control strategies, which includes cardioversion, AADs, and CA. Surgical and atrioventricular node ablation were excluded. The focus of the review was specifically on race disparities in the United States of America. The search was performed using three databases: PubMed/MEDLINE, Web of Science and Embase. We queried the databases from February to March 2024. No time restrictions were placed on publication year in order to capture disparities over time with the evolution of CA indications. We searched the databases using the terms “Atrial Fibrillation” and “Catheter Ablation” or “Rhythm Control” with permutations of race and ethnic categories identified by the Census Bureau American Community Survey (Supplement 2).

### Inclusion and Exclusion Criteria

To be included in the review, papers had to report empirical research and contain quantitative data on the association between race/ethnicity and utilization of CA and rhythm control in atrial fibrillation. We applied the following additional inclusion/exclusion criteria: English language, United States of America as study place of origin, and observational study designs (cross-sectional, retrospective cohorts, and prospective cohorts). The studies had to include self-reported race or ethnicity as a primary exposure variable, with the utilization of CA (excluding surgical and atrioventricular node ablation) and rhythm control strategies (defined as CA, anti-arrhythmic drug use, and electrical cardioversion) as outcome variables. Many data sources are aggregated and may not discuss how race or ethnicity data was categorized or collected. Review articles, commentaries, case reports, case series and letters to the editor were excluded.

### Screening and Review

Search results were imported into a reference manager, and duplicates were removed using an automated tool. The online search yielded 453 results, which were narrowed to 369 articles after removal of duplicates. After titles and abstracts were screened by one reviewer, a total of 32 articles were selected for full-text review. The full text of each reference was then screened for its inclusion eligibility by two independent reviewers (WN and JS). Disagreements were resolved by discussion and consensus between the two reviewers. After applying inclusion/exclusion criteria to the full-text articles, the 32 articles were narrowed to 20. The main reasons for exclusion during the full-text review were article type (e.g. narrative review article) and papers focusing on different exposure or outcome variables (Figure 1).

**Figure 1.**
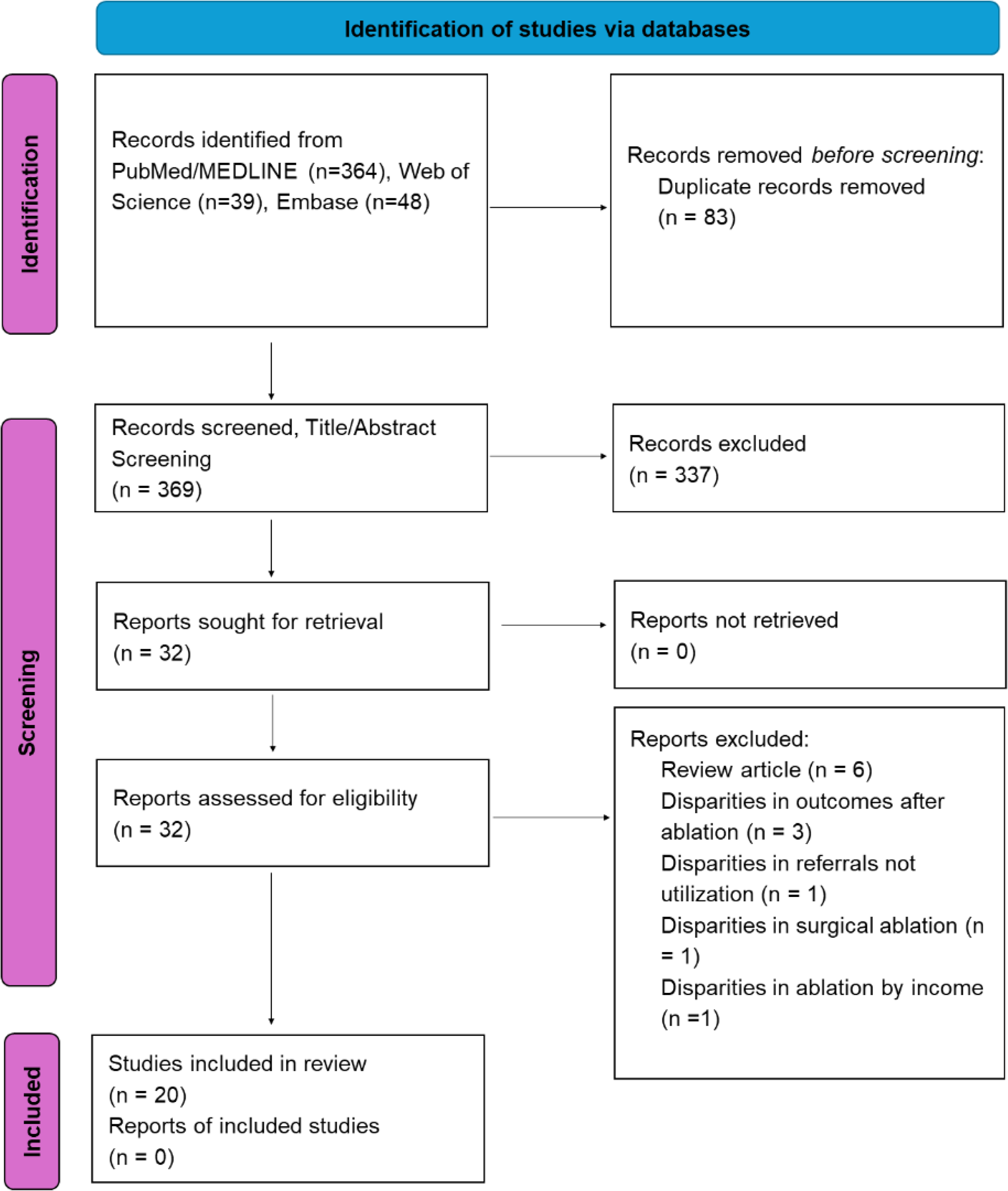
The PRISMA flow diagram for the systematic review detailing the database searches, the number of abstracts screened, and the full texts retrieved.

### Data Extraction

Two reviewers extracted data from the studies included in the review. Variables included study type (i.e., cross-sectional versus longitudinal study design), source of data (i.e., Electronic Health Record [EHR], Administrative/Claims, Registry), study cohort years, sample size, study setting (i.e., inpatient, outpatient, or both), race or ethnic groups included in the study, and patient-level (i.e., demographics, income, insurance type, comorbidities, and measures of community deprivation) and practice-level (i.e., provider type, practice rurality, teach vs non-teach hospital) covariates. We extracted raw data, unadjusted odds ratios, adjusted odds ratio, and p-values, where available, for utilization of CA by race/ethnic group compared to NHW as the reference group.

### Synthesis and Meta-Analysis

Five of 20 studies were included in the narrative review, but were excluded from the meta-analysis due to insufficient data: one study reported utilization by race or ethnic group as a percentage of total ablation procedures with no raw data or effect measures, two studies compared NHW to all other races grouped under “other”, one study only reported population adjusted incidence of ablation by year, and one study was excluded as ablation accounted for <1% of the composite outcome of rhythm control. We performed descriptive analysis to summarize the characteristics of all included studies.

Using unadjusted and adjusted odds and hazards ratios, we calculated a pooled estimate of the utilization of CA by race or ethnicity. Standard errors were estimated from reported 95% confidence intervals and intervals were re-estimated by normal approximation. We calculated and included an unadjusted odds ratio for one study, as only raw data was reported (4). To account for study heterogeneity, we utilized the random effects model of DerSimonian and Laird (8). To assess for publication bias, we performed an Egger’s regression test and produced a funnel plot. A bias coefficient was calculated to confirm the findings of the funnel plot. We also examined between-study heterogeneity using the I2 statistic and prediction intervals.

Analyses were performed using R (Version 4.3.1; R Core Team 2023), RStudio (RStudio Team, 2023) and Stata v. 18 (Stata Corp, College Station, TX).

## Results

This review included 20 studies published between the years 2011 and 2023 (Table 1). The most common reasons for exclusion were review article type (n=6), disparities in outcomes after ablation (n=3), disparities in referrals (n=1), disparities in surgical ablation (n=1) and disparities in ablation by income (n=1). The studies captured 47,700,642 patients with atrial fibrillation, of whom over 561,490 underwent ablation. Studies were published between 2014 and 2023 with cohort years spanning 2000 and 2022. The most common data source type was administrative/claims (n=9), followed by registry (n=6), and electronic health records (EHR) (n=5). Studies mostly derived data from the inpatient setting (n=9), followed by combined inpatients and outpatients (n=6), and outpatients (n=5). The most common study design was longitudinal (n=19). NHB patients were the most represented minority group in studies (n=17), followed by HLx (n=13), Asian (n=9), American Indian/Alaskan Native (n=3) and Native Hawaiian or Other Pacific Islander (n=2). Eight studies included an unspecified race group “other”. Most studies adjusted for demographics such as age (n=16) and sex (n=14), and comorbidities (n=16). Some studies adjusted for practice-level factors (n=11), insurance (n=11) and income (n=5). Only one study adjusted for a neighborhood-deprivation index.

**Table 1.** Descriptive analysis of studies reporting associations between race and utilization of catheter ablation/rhythm control strategies.

| **Variable** | **Groups** | **Number of studies reporting on variable (%)** |
| --- | --- | --- |
| ***Total number of papers*** |  | 20 (100) |
| ***Study Type*** | *Cross-sectional* | 1 (5) |
|  | *Longitudinal* | 19 (95) |
| ***Data Source*** | *Administrative/Claims* | 9 (45) |
|  | *Registry* | 6 (30) |
|  | *EHR* | 5 (25) |
| ***Sample Size*** | *0-9999* | 5 (25) |
|  | *10-99999* | 3 (15) |
|  | *100000-499999* | 4 (20) |
|  | *500000-1000000* | 4 (20) |
|  | *>10000000* | 4 (20) |
| ***Study Setting*** | *Inpatient* | 9 (45) |
|  | *Outpatient* | 5 (25) |
|  | *Both* | 6 (30) |
| ***Race/Ethnic group*** | *Non-Hispanic Black* | 17 (85) |
|  | *Hispanic/LatinX* | 13 (65) |
|  | *Asian* | 9 (45) |
|  | *American Indian or Alaska Native* | 3 (15) |
|  | *Native Hawaiian or Other Pacific Islander* | 2 (10) |
|  | *Other* | 8 (40) |
| *Covariates* | *Age* | 16 (80) |
|  | *Sex* | 14 (70) |
|  | *Comorbidities* | 16 (80) |
|  | *Income* | 5 (25) |
|  | *Insurance* | 11 (55) |
|  | *Distressed Communities Index or other Social Determinant of Health* | 1 (5) |
|  | *Provider or Practice Factors* | 11 (55) |

Some studies reported ablation as part of a composite of rhythm control strategy that also included anti-arrhythmic drug use, and cardioversion (Table 2). Six studies reported an adjusted odds ratio with 95% confidence interval limits and p-values, five studies reported unadjusted and adjusted odds ratios or hazard ratios with 95% confidence interval limits, and four studies reported all relevant data including raw data, unadjusted odds ratios, and adjusted odds ratios with 95% confidence interval limits and p-values. One study only reported raw counts. Two studies reported effect measures by sex; if no combined odds ratio was available, we used reported odds ratios of each sex for the meta-analysis.

**Table 2.** Summary table of studies included in the review.

| **Study** | **Data Source (Year Range)** | **Study Setting** | **Study Method** | **Race or Ethnic Groups Included** | **Covariates Adjusted for in Models** | **Odds of CA ablation by Race or Ethnic Group Compared to NHW (95% CI)** |
| --- | --- | --- | --- | --- | --- | --- |
| **Bhatia et al. 2016 (9)** | Administrative/Claims (2001-2011) | Inpatient | Longitudinal | NHW, NHB, HLx, Asian, Other | Demographics, comorbidities, income, practice factors | NHB aOR 0.830 (0.790-0.880)<br>HLx aOR 0.780 (0.730-0.840)<br>Asian aOR 0.380(0.310-0.450)<br>Other aOR 1.190 (1.080-1.310) |
| **Behave et al. 2015(10)** | Administrative/Claims (2009-2012) | Both | Longitudinal | NHW, NHB, HLx | Demographics, comorbidities, insurance | NHB OR 0.69(0.63-0.76)<br>NHB aOR 1.010(0.910-1.110)<br>HLx OR 0.560 (0.510-0.63)<br>HLx aOR 0.70 (0.63-0.79) |
| **Desai et al. 2021(11)** | Registry (2013-2017) | Inpatient | Longitudinal | NHW, Other | Demographics, comorbidities, practice factors | NHW vs Other* aOR 1.21 (1.08-1.35) |
| **Duke et al. 2022(12)** | EHR (2011-2020) | Outpatient | Longitudinal | NHW, NHB | Demographics, comorbidities, insurance, distressed communities | NHB OR 0.610 (0.410-0.910)<br>NHB aOR 0.630 (0.400-0.980) |
| **Durrani et al. 2018(13)** | EHR (2006-2014) | Both | Longitudinal | NHW, NHB | Demographics, comorbidities | NHB OR 0.370 (0.220-0.800)<br>NHB aOR 0.500(0.100-0.700) |
| **Eberly et al. 2021(14)** | Administrative/Claims (2015-2019) | Both | Longitudinal | NHW, NHB, Asian | Demographics, comorbidities, income, insurance, practice factors | NHB aOR 0.860 (0.730-1.020)<br>HLx aOR 0.730 (0.600-0.890) |
| **Gehi et al. 2017(15)** | Registry (2008-2014) | Outpatient | Longitudinal | NHW, NHB, HLx, Asian, NIAN, NHPI, Other | Demographics, comorbidities, insurance, practice factors | NHW vs Other* aOR 2.429 (2.385-2.473) |
| **Golwala et al. 2016(4)** | Registry (2010-2011) | Outpatient | Longitudinal | NHW, NHB, HLx | None | NHB OR 0.592 (0.409-0.827)<br>HLx OR 0.437 (0.274- |
|  |  |  |  |  |  | 0.658) |
| **Gu et al. 2021(16)** | Registry (2013-2018) | Outpatient | Longitudinal | NHW, Asian | Demographics, comorbidities, practice factors | Asian aOR 0.81 (0.72–0.91) |
| **Hamade et al. 2023(17)** | EHR (2022) | Both | Cross-sectional | NHW, NHB, Other | Demographics, insurance | NHB aOR 0.483 (0.465-0.502)<br>Other aOR: 0.343 (0.332-0.355) |
| **Hoyt et al. 2011(18)** | EHR (2001-2009) | Inpatient | Longitudinal | NHW, NHB, HLx, Asian | None | NA |
| **Jackson et al. 2023(19)** | Administrative/Claims (2011-2018) | Both | Longitudinal | NHW, NHB, HLx, Asian | Demographics, comorbidities, insurance, practice factors | NHB aOR 0.800(0.680-0.940)<br>HLx aOR 0.890 (0.760-1.040)<br>Asian aOR 0.960 (0.740-1.260) |
| **Khalid et al. 2020(20)** | Registry (2008-2016) | Outpatient | Longitudinal | NHW, NIAN | Demographics, comorbidities, practice factors | NIAN<sup>a</sup> OR 0.90 (0.81 to 0.99) |
| **Kir et al. 2022<sup>b</sup>(21)** | Registry (2013-2019) | Inpatient | Longitudinal | NHW, NHB, HLx, Asian, Other | Demographics, comorbidities, insurance, practice factors | NHB aOR 0.849 (0.651-1.108)<br>HLx aOR 0.614(0.439-0.859)<br>Asian aOR 1.137 (0.745-1.736)<br>Other aOR 0.511 (0.347-0.753) p=0.001 |
| **Kummer et al. 2015(22)** | Administrative/Claims (2005-2011) | Inpatient | Longitudinal | NHW, NHB, HLx, Asian, Other | Demographics, Comorbidities, Income, Insurance, | NHB AB aHR 0.680 (0.630-0.730)<br>HLx aHR 0.600 (0.560-0.640)<br>Asian aHR 0.580 (0.510-0.660)<br>Other aHR 0.770 (0.690-0.860) |
| **Naderi et al. 2014(23)** | Administrative/Claims (2009-2010) | Inpatient | Longitudinal | NHW, NHB | Demographics, Comorbidities | NHB aOR (males) 0.80 (0.72-0.88)<br>Other aOR (male) 0.70 (0.65-0.77)<br>NHB aOR (females) 0.44 (0.39-0.50)<br>Other aOR (female) 0.38 (0.34-.043) |
| **Patel et al. 2016(24)** | Administrative/Claims (2000-2012) | Inpatient | Longitudinal | NHW, NHB, HLx, Other | Demographics, comorbidities, insurance, income, practice factors | NHB aOR 0.49(0.44-0.55)<br>HLx aOR 0.64(0.56-0.72)<br>Others aOR 0.92(0.82-1.03) |
| **Tamariz et al. 2014(25)** | Administrative/Claims (2006-2009) | Both | Longitudinal | NHW, NHB | Demographics, Comorbidities, Insurance, Practice Factors | NHB OR 0.590 (0.520-0.660)<br>NHB aOR 0.670(0.600-0.750)<br>HLx OR: 0.820 (0.740- |
|  |  |  |  |  |  | 0.900)<br>HLX aOR: 0.830 (0.750-0.910) |
| **Tien et al. 2023(26)** | EHR<br>(2019-2022) | Inpatient | Longitudinal | NHW,<br>NHB,<br>HLx | None | NA |
| **Yarrarapu et al. 2023 (27)** | Administrative/Claims<br>(2007-2017) | Inpatient | Longitudinal | NHW,<br>NHB,<br>HLx,<br>Asian,<br>NIAN,<br>NHPI | Demographics,<br>comorbidities,<br>income,<br>insurance,<br>practice factors | NHB aOR 0.695 (0.674-0.718)<br>HLx aOR 0.788 (0.759-0.819)<br>Asian aOR 0.609 (0.568-0.652)<br>NIAN aOR 1.39 (1.245-1.552) |
| *Other included includes non-Hispanic Black, Asian, American Indian/Alaskan Native, Native Hawaiian/Pacific Islander, mixed, and missing patients.<br><sup>a</sup>Reported as a composite outcome of rhythm control strategies<br><sup>b</sup>Catheter ablation made up <1% of the composite out of rhythm control. |  |  |  |  |  |  |

Compared to NHW patients, NHB patients had a lower likelihood of utilizing CA for atrial fibrillation, pooled OR of 0.68 (95% CI [0.58-0.77], k = 14). Hispanic/Latinx (HLx) and Asian patients were also shown to have a lower likelihood of utilization CA, pooled OR of 0.72 (95% CI [0.65-0.79], k = 9) and OR of 0.62 (95% CI [0.45-0.78], k = 5) respectively. We generated a forest plot of the studies included in the analysis with weighted effect sizes (Figure 2).

**Figure 2.**
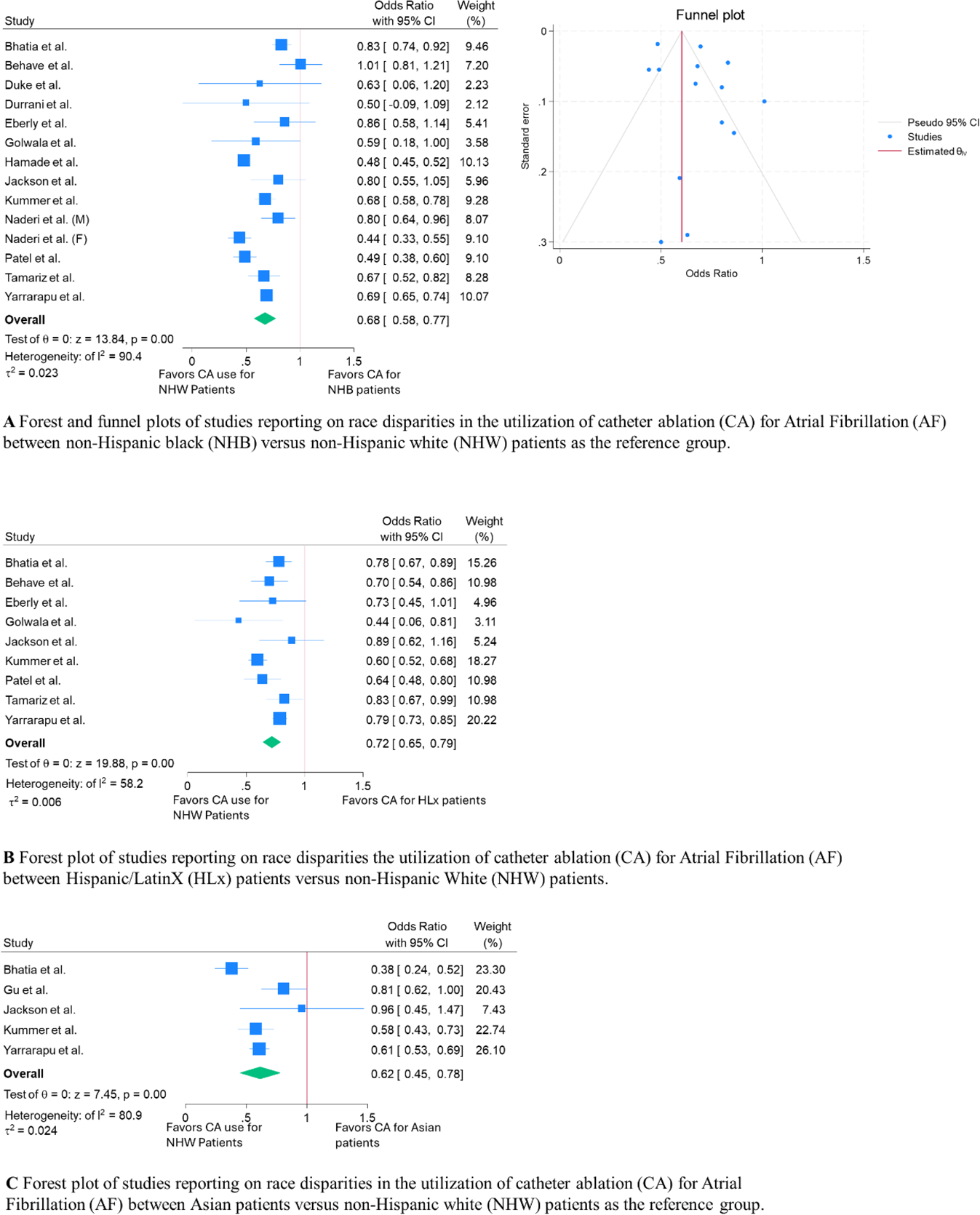
Race and ethnic disparities in the utilization of catheter ablation for AF. Forest plots based on a random effects model of included studies by race/ethnicity: non-Hispanic Black patients (A) reported with a funnel plot, Hispanic/LatinX patients (B) and Asian patients (C). A square represents the binary effect size reported by each study. The size of the box is determined by the study weight. The diamond represents pooled effect estimate. I^2^ and τ^2^ values are reported.

Only the estimate for NHB patients was pooled from enough studies (>10) to assess publication bias through a funnel plot (figure 2A) which showed asymmetry. An I^2^ value was generated by race group: HLx (I^2^ = 58.2%), Asian (I^2^ = 80.9%), and NHB (I^2^ = 90.4%).

## Discussion

The pooled ORs of 0.68 (95%CI 0.58-0.77) for non-Hispanic Black, 0.72 (95%CI 0.65-0.79) for Hispanic/LatinX, and 0.62 (95%CI 0.45-0.78) for Asian patients reveal disparities in CA utilization for AF among minority groups compared NHW. However, the high degree of between-study heterogeneity noted by the meta-analysis demonstrates that no single pooled estimate can summarize these disparities, but rather points to varying degrees of disparities across different populations and settings that requires further explanation. Nonetheless, our results show persistent race/ethnic disparities in the utilization of CA for AF that span several years and both the inpatient and outpatient settings.

These disparities persisted despite some studies adjusting for multiple covariates including demographics, income, insurance, comorbidities, and provider or practice factors. Of note, certain ethnic/race groups including American Indians/Native Alaskans and Native Hawaiians/other Pacific Islanders were underrepresented in the disparities literature, limiting their inclusion in the meta-analysis portion of the review. Our findings are consistent with the literature which shows disparities in the use of cardiac procedures in the United States such as diagnostic and therapeutic catheterization in ischemic heart disease, trans-aortic and surgical aortic valve replacement, and implantable cardioverter-defibrillator for primary prevention sudden cardiac death (28–30).

Given the higher morbidity and mortality of AF in minority groups and recent emphasis in national guidelines on early rhythm control, understanding the root causes of race disparities in CA is critical to ensuring equitable access to this important intervention. Several non-clinical and clinical factors may contribute to disparities in the utilization of health services such as CA. To better conceptualize these factors, we adapted the Andersen’s behavioral model (ABM) of health services utilization, an established model that has been applied to understand factors that facilitate or impede health services use in a broad range of patient populations and disease states (31,32).

The ABM predicts that an individual’s use of a health service is a function of three elements: (1) one’s predisposition to use of a service; (2) factors that enable the use of a service; and (3) the need for a service, which may include an individual’s perceived need and their physician’s judgement and assessment of need (Figure 3). Unlike other health behavioral models of service utilization, the ABM considers the interaction of these elements at both the individual and environmental level to predict service utilization. We explore how predisposing, enabling and need factors at the individual and environmental level may explain the persistent race/ethnic disparities in the utilization of catheter ablation utilization for atrial fibrillation and propose areas for further research to clarify their role in disparities (Table 3).

**Figure 3.**
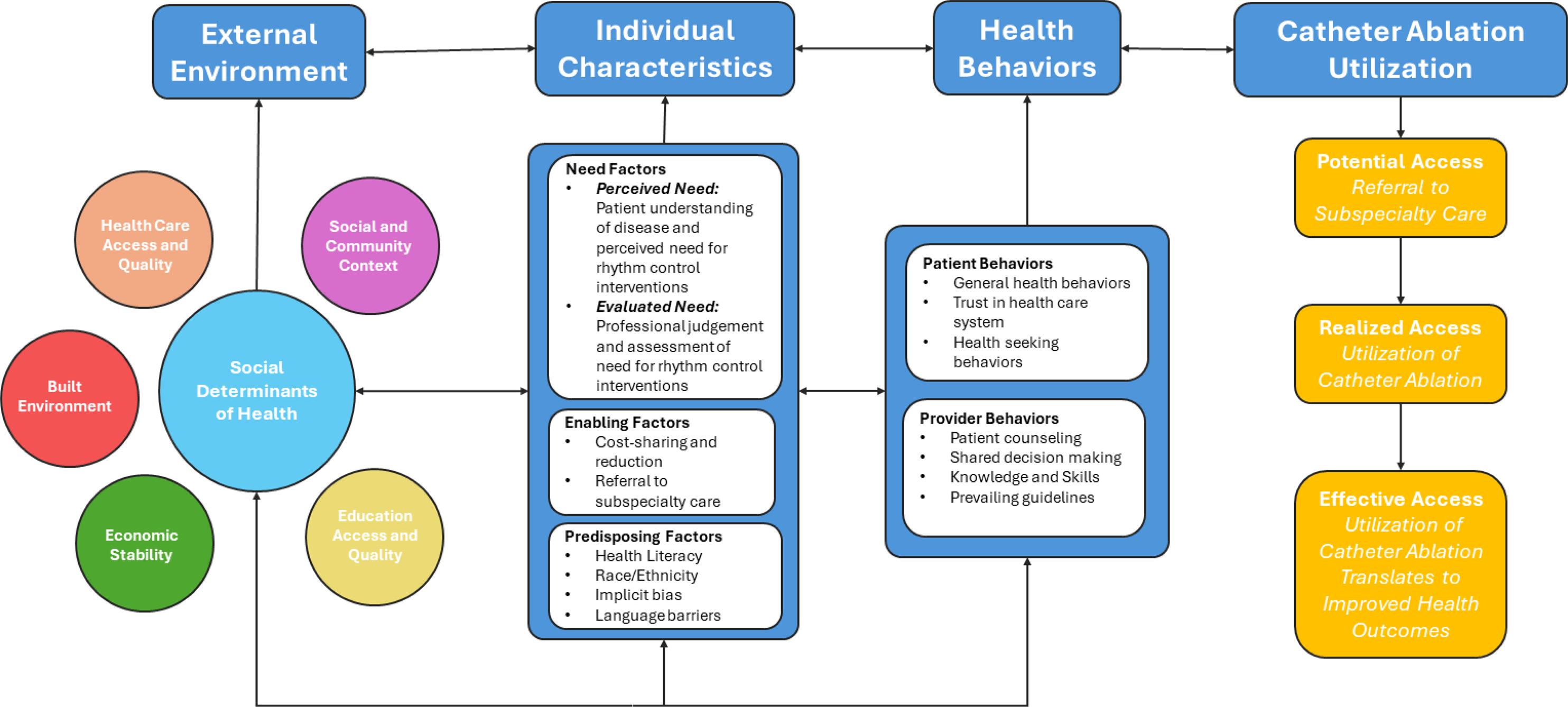
Adapting the Andersen behavioral model of health services utilization, we highlight the interactions between environmental factors (left of the diagram) and individual factors and health behaviors (middle of diagram) which may impede or promote utilization of catheter ablation (right of diagram).

**Table 3.**
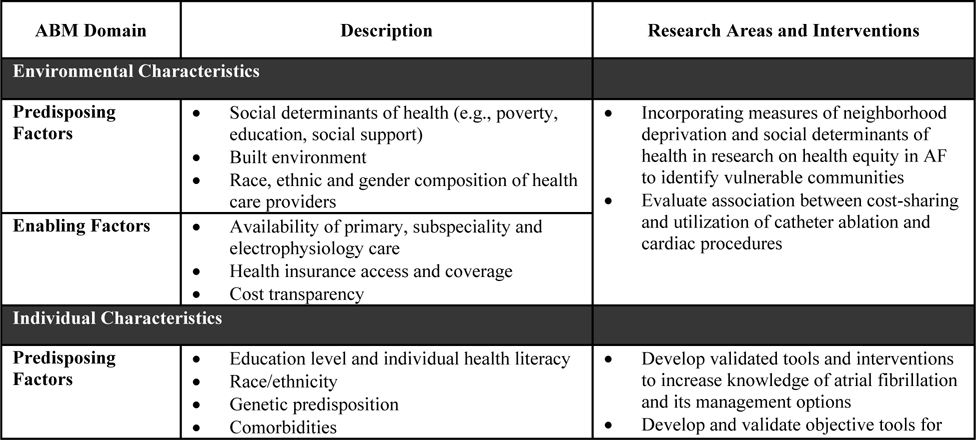
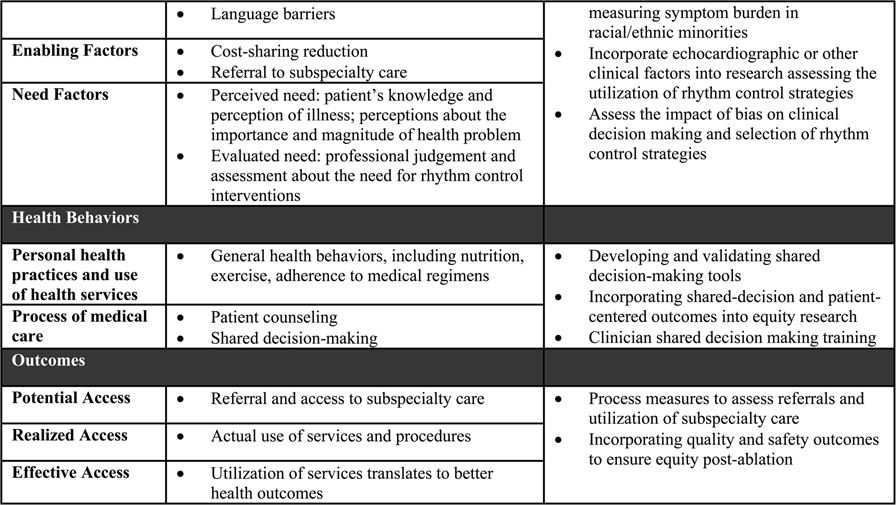
Andersen’s Behavioral Model (ABM) of health services use domains with description of individual and environmental characteristics, and potential areas for implementation research.

The association between environmental factors and utilization of AF care has been documented previously, with one study showing an association between measures of neighborhood deprivation and the likelihood of receiving interventions like CA(12). The study revealed that NHB patients were more likely to come from materially deprived communities as measured by the Distressed Community Index based on US Census Bureau data (i.e., the index combines metrics on education level, housing vacancy rate, percent unemployment, percent living under the poverty line, median income, change in employment and change in business establishments). Yet disparities in ablation persisted even after adjusting for this variable, suggesting that simple zip-code-level distress scores may not sufficiently adjust for or capture important social determinants of health such as transportation, opportunity costs and ability to take time off work, and other non-work obligations that may impede utilization. Incentives, policies, and systems are needed to collect more granular data on social determinants of health to clarify their role in perpetuating disparities in CA utilization.

Insurance type, namely Medicaid or uninsured status, has been associated with lower likelihood of undergoing CA compared to patients who are dual enrolled in Medicare or have private insurance(17). However, multiple studies in our review showed persistent race disparities after adjusting for insurance type (24,25). These findings highlight the multidimensional nature of health insurance coverage, where utilization is not only determined by access alone but also the scope of benefits and the breadth of coverage including out-of-pocket costs. For example, reduction of out-of-pocket costs for medications in post-myocardial infarction patients in the Post-Myocardial Infarction Free Rx Event and Economic Evaluation (MI FREEE) trial was shown to improve adherence to therapy among non-white patients and reduce race and ethnic disparities in clinical outcomes (33). Further attention must be directed towards exploring how cost-sharing and price transparency may affect disparities in CA utilization.

Among various studies included in our review, race disparities remained also after adjusting for individual factors at the patient and provider level. Although minority patients often have higher rates of comorbidities, they still had a lower likelihood of CA despite adjusting for comorbidities. However, observed disparities may be secondary to unmeasured clinical factors which are not captured by the data sources used by studies such as EHRs and administrative/claims data. Notably, because these data are derived from ICD codes, there may be errors in coding as well as a lack of important information on disease severity. Other important clinical data such as echocardiographic parameters, which may influence decisions about pursuing ablation or rhythm control, were also missing. Future research should examine whether disparities despite after adjusting for echocardiographic data and disease severity. Other notable patient factors, like symptom burden, were also not captured by data sources used for the studies in our review despite symptom burden being key driver in determining whether to pursue CA. Objective measures of symptom burden, including validated symptom scales and rhythm monitors may help to clarify whether observed disparities are partially explained by differences in the assessment of symptoms or AF burden by patients and providers.

Race and ethnic disparities may also be mediated by unmeasured provider or practice factors. The data presented by Gehi et al. based on an outpatient registry showed a large variation in approaches to rhythm control across practices, with some practices pursuing rhythm control in as little as <5% of patients while others pursuing rhythm in >50% of patients (15). Using reference effect measures methodology, unmeasured practice factors appeared to contribute more to this wide variation in rhythm control utilization compared to measured patient and practice factors in their model such as race, comorbidities, insurance type and whether the patient was seen by an electrophysiology physician. Unmeasured and potentially modifiable practice factors, which may be contributing to disparities, should be identified and eliminated to standardize care across practice settings.

The relationship between need factors, as identified by the ABM model, and disparities in CA utilization were not examined in any of the studies included in our review. Need factors, including perceived need for an intervention by the patient and evaluated need as assessed by the clinician, ultimately determine whether a patient is motivated to utilize care (Table 3). A patient’s perceived need for CA may be heavily influenced by disease knowledge and health literacy while a physician’s assessment of the patient’s need for a procedure requires the integration of clinical data, patient suitability, and practice guidelines.

Studies have shown that shared decision making (SDM) may enhance patient disease knowledge and willingness to participate in discussions about disease management with their physician in the field of cardiology (34). Research on SDM and health literacy remains limited in AF care, despite being mandated as a priority for research by professional societies (35,36). For example, among patients with new-onset AF in a national registry, only one in four reported that their decision on anticoagulation was the result of SDM with their physician and one in five reported that they participated in SDM when choosing their rhythm control strategy (37). While SDM in AF care appears to be poorly implemented globally, race and ethnic disparities in SDM and patient literacy should be further investigated to assess whether they contribute disparities in the utilization of CA.

The ABM model recognizes that evaluated need by a physician not only represents objective measurements about a patient’s need for a therapeutic intervention, but also has a social component that varies with evolving practice patterns, clinical inertia, implicit bias, and the training background and competency of the professional performing the assessment (32). Implicit racial/ethnic bias is an important social factor not traditionally included in the model but may contribute to disparities in adopting rhythm control or CA. Implicit bias, not only with regards to race but gender and other biases, has been shown to be present among health care workers and may contribute to lower quality of care (38).

There are several limitations of this review. One major limitation is that our search was restricted to databases available at our institution and may potentially exclude studies only reported in other databases or the grey literature. There was also no formal assessment of individual study quality and bias. Although we attempted to include studies that adjusted for similar covariates, there remained a large degree of heterogeneity in studies which we attempted to account for using a random effects model for the meta-analysis. High degree of heterogeneity is to be expected when assessing disparities in utilization over time as our studies differed in a key number of areas including sample size, types of populations sampled, years data are collected and covariate adjustments. Finally, our model assumes that the studies included are sampled from a universal population that approximates the normal distribution, which otherwise may not be true and could affect our pooled estimates. We also detected publication bias based on a funnel plot for NHB patients, which may result in overestimation of the disparities noted in the study. We were unable to assess publication bias for other race groups due to insufficient data.

## Conclusions

Overall, our review indicates the presence of persistent health disparities in the utilization of CA over time even after studies in the literature adjusted for important covariates. By applying a commonly used behavioral model of health service utilization, we identify several individual and environmental factors which may contribute to disparities but are not captured by current studies which rely on EHR or administrative data. Further research adjusting for patient-provider preferences and factors, echocardiographic data and social determinants of health is needed to clarify root causes of disparities and to promote equitable adoption of this important therapy in AF care. Future research should include other minority groups who were underrepresented in the disparities literature including Asian Americans, American Indians, Native Alaskans and Native Hawaiians or Pacific Islanders.

## Abbreviations

AF: Atrial fibrillation
CA: catheter ablation
NHW: non-Hispanic White
NHB: non-Hispanic Black
HLx: Hispanic/LatinX
AAD: anti-arrhythmic drugs
OR: odds ratio
HR: hazard ratio
ABM: Anderesen Behavioral Model
SDM: shared decision making

## Data Availability

Supplement material will be provided with submission.

## References

1. Martin SS, Aday AW, Almarzooq ZI et al. 2024 Heart Disease and Stroke Statistics: A Report of US and Global Data From the American Heart Association. Circulation 2024;149:e347–e913.

2. Ugowe FE, Jackson LR, 2nd, Thomas KL. Racial and ethnic differences in the prevalence, management, and outcomes in patients with atrial fibrillation: A systematic review. Heart Rhythm 2018;15:1337–1345.

3. Tamirisa KP, Al-Khatib SM, Mohanty S et al. Racial and Ethnic Differences in the Management of Atrial Fibrillation. CJC Open 2021;3:S137–S148.

4. Golwala H, Jackson LR, 2nd, Simon DN et al. Racial/ethnic differences in atrial fibrillation symptoms, treatment patterns, and outcomes: Insights from Outcomes Registry for Better Informed Treatment for Atrial Fibrillation Registry. Am Heart J 2016;174:29–36.

5. Magnani JW, Norby FL, Agarwal SK et al. Racial Differences in Atrial Fibrillation-Related Cardiovascular Disease and Mortality: The Atherosclerosis Risk in Communities (ARIC) Study. JAMA Cardiology 2016;1:433–441.

6. Camm AJ, Naccarelli GV, Mittal S et al. The Increasing Role of Rhythm Control in Patients With Atrial Fibrillation. Journal of the American College of Cardiology 2022;79:1932–1948.

7. Joglar JA, Chung MK, Armbruster AL et al. 2023 ACC/AHA/ACCP/HRS Guideline for the Diagnosis and Management of Atrial Fibrillation: A Report of the American College of Cardiology/American Heart Association Joint Committee on Clinical Practice Guidelines. Circulation 2024;149:e1–e156.

8. DerSimonian R, Laird N. Meta-analysis in clinical trials. Controlled Clinical Trials 1986;7:177–188.

9. Bhatia S, Qazi M, Erande A et al. Racial Differences in the Prevalence and Outcomes of Atrial Fibrillation in Patients Hospitalized With Heart Failure. Am J Cardiol 2016;117:1468–73.

10. Bhave PD, Lu X, Girotra S, Kamel H, Vaughan Sarrazin MS. Race- and sex-related differences in care for patients newly diagnosed with atrial fibrillation. Heart Rhythm 2015;12:1406–12.

11. Desai NR, Sciria CT, Zhao X et al. Patterns of Care for Atrial Fibrillation Before, During, and at Discharge From Hospitalization. Circulation: Arrhythmia and Electrophysiology 2021;14:e009003.

12. Duke JM, Muhammad LN, Song J et al. Racial Disparity in Referral for Catheter Ablation for Atrial Fibrillation at a Single Integrated Health System. J Am Heart Assoc 2022;11:e025831.

13. Durrani AF, Soma S, Althouse AD, Leef G, Qin D, Saba S. Impact of Race on Outcome of Patients Undergoing Rhythm Control of Atrial Fibrillation. J Immigr Minor Health 2018;20:14–19.

14. Eberly LA, Garg L, Yang L et al. Racial/Ethnic and Socioeconomic Disparities in Management of Incident Paroxysmal Atrial Fibrillation. JAMA Netw Open 2021;4:e210247.

15. Gehi AK, Doros G, Glorioso TJ et al. Factors associated with rhythm control treatment decisions in patients with atrial fibrillation-Insights from the NCDR PINNACLE registry. Am Heart J 2017;187:88–97.

16. Gu K, Mahtta D, Kaneria A et al. Racial disparities among Asian Americans with atrial fibrillation: An analysis from the NCDR® PINNACLE Registry. Int J Cardiol 2021;329:209–216.

17. Hamade H, Jabri A, Mishra P, Butt MU, Sallam S, Karim S. Gender, ethnic, and socioeconomic differences in access to catheter ablation therapy in patients with atrial fibrillation. FRONTIERS IN CARDIOVASCULAR MEDICINE 2023;9.

18. Hoyt H, Nazarian S, Alhumaid F et al. Demographic profile of patients undergoing catheter ablation of atrial fibrillation. J Cardiovasc Electrophysiol 2011;22:994–8.

19. Jackson LR, Friedman DJ, Francis DM et al. Race and Ethnic and Sex Differences in Rhythm Control Treatment of Incident Atrial Fibrillation. CLINICOECONOMICS AND OUTCOMES RESEARCH 2023;15:387–395.

20. Khalid U, Marzec LN, Mantini N et al. Treatment of AF in American Indians and Alaska Natives: Insights From the NCDR PINNACLE-AF Registry. Journal of the American College of Cardiology 2020;75:2749–2750.

21. Kir D, Zhang S, Kaltenbach LA et al. Patterns of care for first-detected atrial fibrillation: Insights from the Get With The Guidelines® -Atrial Fibrillation registry. Heart Rhythm 2022;19:1049–1057.

22. Kummer BR, Bhave PD, Merkler AE, Gialdini G, Okin PM, Kamel H. Demographic Differences in Catheter Ablation After Hospital Presentation With Symptomatic Atrial Fibrillation. J Am Heart Assoc 2015;4:e002097.

23. Naderi S, Rodriguez F, Wang Y, Foody JM. Racial disparities in hospitalizations, procedural treatments and mortality of patients hospitalized with atrial fibrillation. Ethn Dis 2014;24:144–9.

24. Patel N, Deshmukh A, Thakkar B, et al. Gender, Race, and Health Insurance Status in Patients Undergoing Catheter Ablation for Atrial Fibrillation. Am J Cardiol 2016;117:1117–26.

25. Tamariz L, Rodriguez A, Palacio A, Li H, Myerburg R. Racial disparities in the use of catheter ablation for atrial fibrillation and flutter. Clin Cardiol 2014;37:733–7.

26. Tien M, Saddic LA, Neelankavil JP, Shemin RJ, Williams TM. The Impact of COVID-19 on Racial and Ethnic Disparities in Cardiac Procedural Care. J Cardiothorac Vasc Anesth 2023;37:732–747.

27. Yarrarapu SNS, Shah P, Iskander B, et al. Epidemiology, Trends, Utilization Disparities, and Outcomes of Catheter Ablation and Its Association With Coronary Vasospasm Amongst Patients With Non-valvular Atrial Fibrillation: A Nationwide Burden of Last Decade. Cureus 2023;15:e40649.

28. Cromwell J, McCall NT, Burton J, Urato C. Race/ethnic disparities in utilization of lifesaving technologies by Medicare ischemic heart disease beneficiaries. Med Care 2005;43:330–7.

29. Patel NJ, Edla S, Deshmukh A, et al. Gender, Racial, and Health Insurance Differences in the Trend of Implantable Cardioverter-Defibrillator (ICD) Utilization: A United States Experience Over the Last Decade. Clin Cardiol 2016;39:63–71.

30. Kulkarni A, Arafat M, Hou L, Liang S, Kassotis J. Racial Disparity Among Patients Undergoing Surgical Aortic Valve Replacement and Transcatheter Aortic Valve Replacement in the United States. Angiology 2023;74:812–821.

31. Mareike L, Jana T, Eva MB. Application of Andersen’s behavioural model of health services use: a scoping review with a focus on qualitative health services research. BMJ Open 2021;11:e045018.

32. Andersen RM, Davidson PL. Improving Access to Care in America: Individual and Contextual Indicators. Changing the US health care system: Key issues in health services policy and management, 3rd ed. San Francisco, CA, US: Jossey-Bass, 2007:3–31.

33. Choudhry NK, Bykov K, Shrank WH et al. Eliminating Medication Copayments Reduces Disparities In Cardiovascular Care. Health Affairs 2014;33:863–870.

34. Panagiota M, Nicolai G-H, Johannes R, Charikleia P. Shared decision making in cardiology: a systematic review and meta-analysis. Heart 2023;109:34.

35. Aronis KN, Edgar B, Lin W, Martins MAP, Paasche-Orlow MK, Magnani JW. Health Literacy and Atrial Fibrillation: Relevance and Future Directions for Patient-centred Care. Eur Cardiol 2017;12:52–7.

36. Magnani JW, Mujahid MS, Aronow HD et al. Health Literacy and Cardiovascular Disease: Fundamental Relevance to Primary and Secondary Prevention: A Scientific Statement From the American Heart Association. Circulation 2018;138:e48–e74.

37. Ali-Ahmed F, Pieper K, North R et al. Shared decision-making in atrial fibrillation: patient-reported involvement in treatment decisions. European Heart Journal - Quality of Care and Clinical Outcomes 2020;6:263–272.

38. FitzGerald C, Hurst S. Implicit bias in healthcare professionals: a systematic review. BMC Medical Ethics 2017;18:19.

